# Health and Economic Costs of Early, Delayed and No Suppression of COVID-19: The Case of Australia

**DOI:** 10.1101/2020.06.21.20136549

**Authors:** Tom Kompas, R. Quentin Grafton, Tuong Nhu Che, Long Chu, James Camac

## Abstract

We compare the health and economic costs of early (actual), delayed and no suppression of COVID-19 infections in 2020 in Australia. Using a fit-for-purpose compartment model that we fitted from recorded data, a value of a statistical life year (VSLY) and an age-adjusted value of statistical life (A-VSL), we find: (1) the economic costs of no suppression are multiples more than for early suppression; (2) VSLY welfare losses of fatalities equivalent to GDP losses mean that for early suppression to *not* to be the preferred strategy requires that Australians prefer more than 12,500–30,000 deaths to the economy costs of early suppression, depending on the fatality rate; and (3) early rather than delayed suppression imposes much lower economy and health costs. We conclude that in high-income countries, like Australia, a ‘go early, go hard’ strategy to suppress COVID-19 results in the lowest estimated public health and economy costs.

## 1. Introduction

As of the end of May 2020, the global number of COVID-19 cumulative cases and reported fatalities, respectively, exceed 6 million and 370,000 (Johns Hopkins University, 2020). In response to the pandemic, many countries have imposed various types of suppression measures at different times to reduce the growth rate in COVID-19 infections (Johns Hopkins University, 2020). Severe suppression measures (e.g., school and university closures; travel restrictions; mandatory social distancing requirements), when implemented in combination, have received the moniker, ‘lock-down’. Some (including the President of the United States on 22 March 2020) have questioned whether the ‘cure’ (i.e., lock-down) may be more costly in terms of the economy than no or limited suppression measures (e.g., Singer, P. and Plant, M. (2020), Frijters (2020), van den Broek-Altenburg, E. and Atherly, A. (2020)). This question can only be resolved with an epidemiological model of COVID-19 infections estimated from actual data combined with an empirical economic model of the costs of a lock-down. Using a combined epi-economic modeling approach we estimate and compare the public health and economic costs of early, late and no suppression measures.

We evaluate whether a lock-down to suppress COVID-19 in Australia was justified by estimating the health and economic costs (including welfare costs of COVID-19 patients) for three scenarios. The first scenario is ‘early suppression’ and is what actually occurred with the imposition of a lock-down that began in March 2020, and that we simulate continued until the end of May 2020. The second is ‘delayed suppression’ or the effect of imposing a lock-down either 14, 21 or 28 days after it was actually imposed. For completeness, the third scenario is a ‘no suppression’ counter-factual that assumes there were no Australian government (and voluntary) suppression measures. For each scenario, we estimate the number of fatalities, hospitalisations, and economic costs.

As of 1 June 2020, Australian suppression measures have drastically reduced community transmission of the virus and the daily growth rate (three-day average) has declined from around 25%, with 268 new recorded cases on 22 March, to a daily growth rate of 0.26% and 11 new recorded cases on 30 May 2020 (Covid19-Data, 2020). The total number of recorded cases and fatalities, respectively, are just over 7,000 and 100. Using Australian data, we analyze the public health effects of early, delayed and no suppression measures from a Susceptible, Infected, Quarantined, Recovered, and Mortality (SIQRM) model we constructed for this purpose. Estimates of the welfare losses for COVID-19 patients are calculated using an Australian value of statistical life year (VSLY) and an age-adjusted value of a statistical life (A-VSL). Our key finding is that the economic costs (including loss of life) of early suppression are much less than delayed or no suppression measures for a large range of possible outcomes.

## 2. COVID-19 in Australia and policy responses

COVID-19 was first detected in Australia in January 25th, 2020, from a passenger who arrived in Melbourne from Wuhan on January 19th. As of May 30th, 2020 there were 7,173 reported cases and of which 6,582 have recovered with 102 deaths (Covid19-Data, 2020). Figure 1 provides a summary of cumulative reported infected cases and the daily reported growth rate (3-day average).

**Figure 1:**
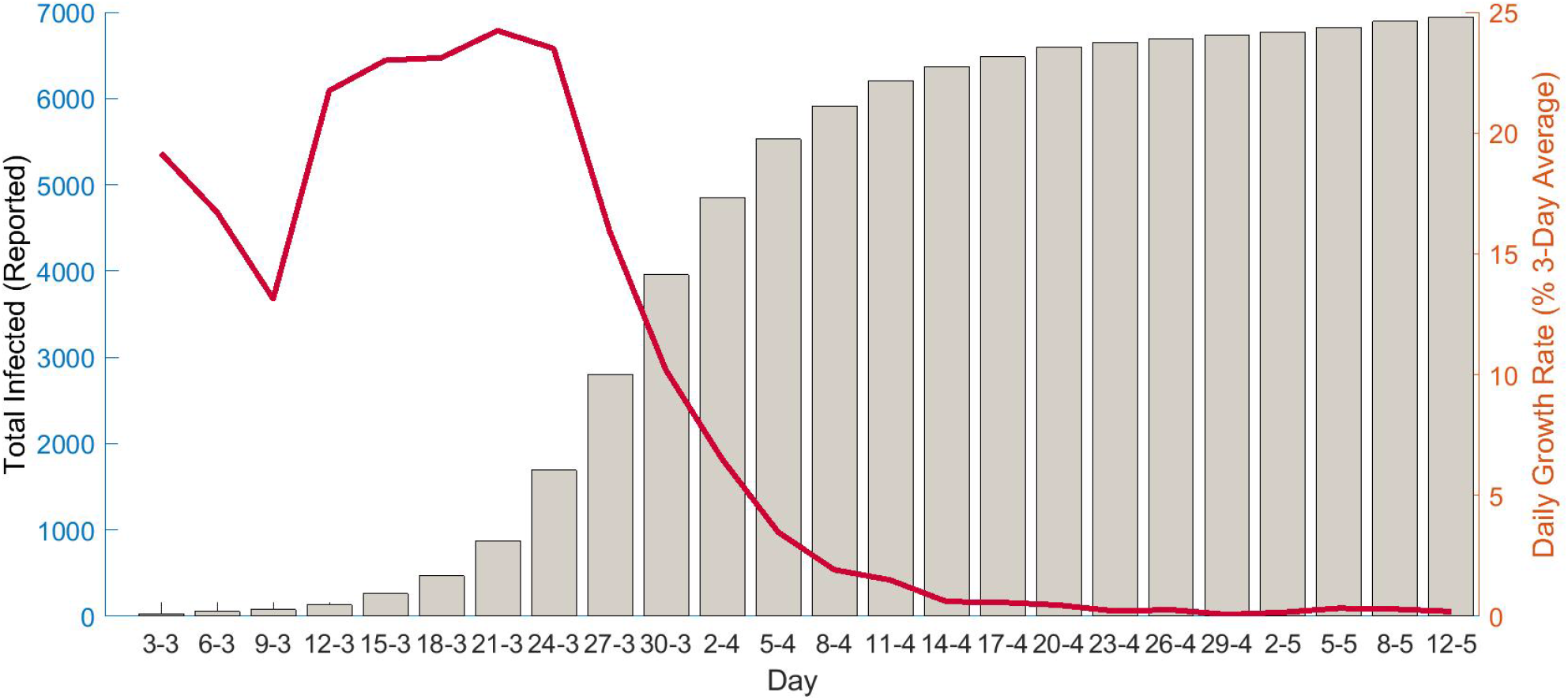
COVID-19 Infected Cases and 3-Day Average Growth Rate. Source: Covid19-Data (2020)

The first recorded death from COVID-19 in Australia was on March 1st, 2020 when the total number of cases in the country was 29. During the initial stages of the infection, the total number of cases (including overseas arrivals) and the number of active cases grew exponentially, which prompted the Australian government to respond with a series of sequential and increasingly strict control measures.

Figure 2 summarizes the dynamics of COVID-19 and the main control responses in Australia since the first Australian death. Control responses varied by state and territory.

**Figure 2:**
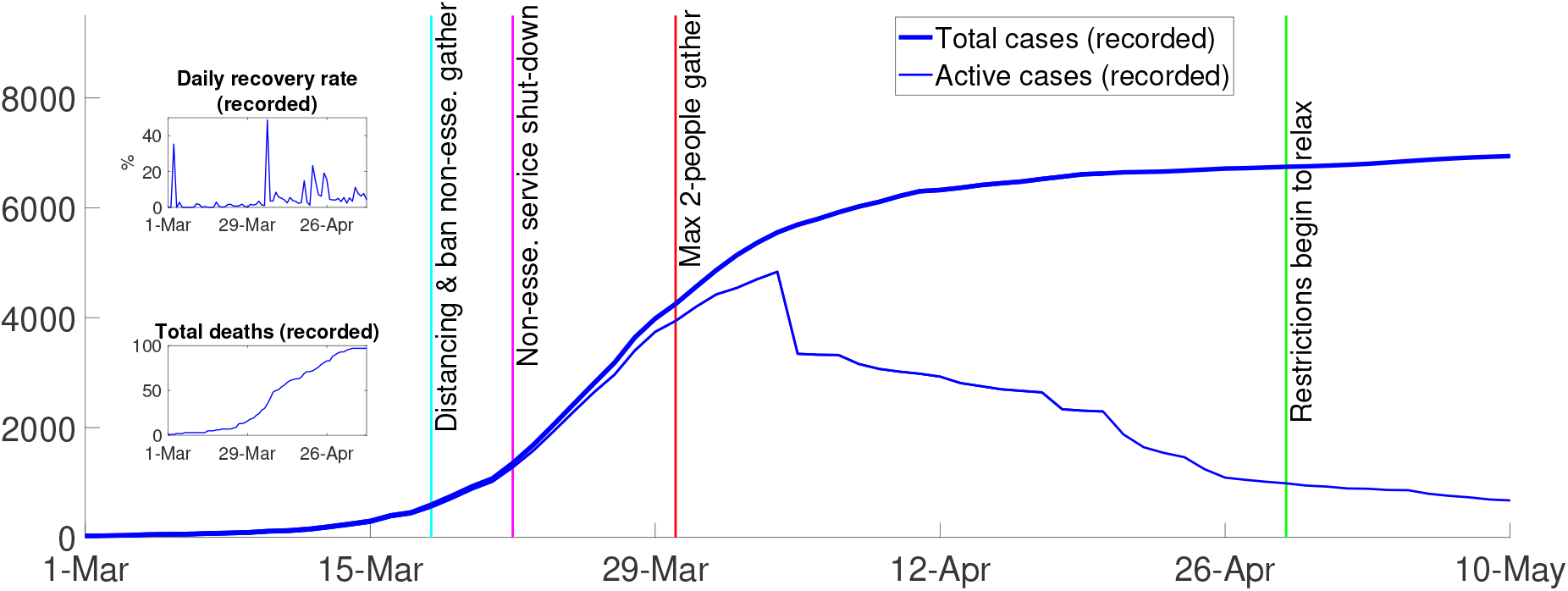
Overview of COVID-19 in Australia. Source: Covid19-Data (2020)

From March 15th, the Australian federal government required international arrivals to self-quarantine for 14 days. Three days later, the government banned non-essential gatherings and encouraged social distancing (i.e., 1-5m distance and 4m2 per person). On March 21st, the federal government ordered non-essential businesses to close. Non-essential businesses included pubs, licensed clubs, hotels, places of worship, gyms, indoor sporting venue, cinemas, and casinos. Some further tightening also occurred, shortly after, with more restrictions on funeral and wedding attendance, fitness classes, and arcades (Australian Department of Health, 2020a).

The most severe suppression measures were introduced on March 30th when public gatherings were reduced to a maximum of two people. From this date, Australians were required to stay at home unless shopping for essentials, receiving medical care, limited (30 minute) exercising, or traveling to and from work or for educational purposes. All suppression measures combined ‘flattened the curve’ and rapidly reduced the number of reported infected cases. Figure 2 shows that recoveries started exceeding new infections on April 4th when the number of recorded active cases peaked at 4,935.

## 3. Methods

### 3.1. Epidemiological model

We constructed an epidemiological model of COVID-19 infections in Australia that has five compartments, **S**usceptible, **I**nfected, **Q**uarantined, **R**ecovered, and **M**ortality, using recorded data for **Q, R** and **M**. Susceptible people can be infected with the virus via community transmission and become infectious. People who are infected can also come from abroad and transmit the disease before clinical signs appear. We assumed, as implemented in Australia, that all those who are officially reported to be infected are ‘quarantined’, recorded as active cases, in hospital or self-isolating, and either recover or die. Suppression measures mitigate the spread of the infection while the speed of recovery depends on treatment protocols.

Our model is formalized as follows:

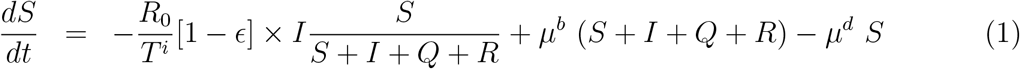

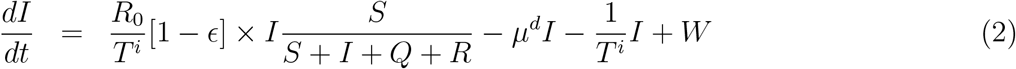

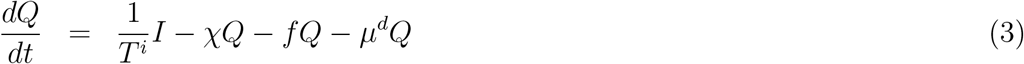

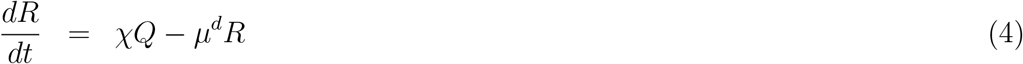

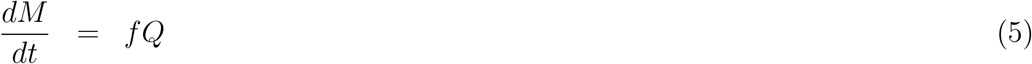

where *S, I, Q, R, M* are the number of susceptible, infected, recorded active cases (i.e., quarantined), and eventually recorded as recovered and deceased, all functions of time; *W* is the number of infected people who arrive from abroad; *R*_0_ is the basic reproduction rate (see Wu et al. (2020)); is the effectiveness of suppression measures; *T* ^*i*^ is the infectious period; *µ*^*d*^ is the natural mortality rate (we estimate *µ*^*d*^ = 0.7% using the mortality data from the Australian Institute of Health and Welfare (2019) and population data from the ABS (2017)); *µ*^*b*^ is the birth rate (we specify *mu*^*b*^ = 1.3% using the estimates from the ABS (2020a)); *χ* is the rate at which active cases recover, which is observable (as in the top subplot in Figure 2); and *f* is the rate at which infected people die because of the virus, where *f* = 1.67%, as given from reported data in late April, and varies daily over the model run from March 1st to April 20th (Covid19-Data, 2020). Economy results are also shown for the fatality rate in Verity et al. (2020) of 0.7%.

The policy variable (*t*) represents suppression measures and is delimited by 0*≤* [1*−∈*(*t*)] *≤* 1, for combined mandated and voluntary measures. Thus, without suppression measures [1*−* ∈ (*t*)] approaches one (i.e., (*t*) approaches 0) and with the most restrictive suppression measures [1*−* ∈ (*t*)] approaches zero (i.e., (*t*) approaches 1).

The total number of susceptible people is assumed to be 70% of the total population and, in effect, excluded the below-20-year-old population group. From the SIQRM model in equations 1-5, the total number of recorded cases (*T*) were calculated as the sum of active cases (in quarantine), recovered, and deaths, or *T* = *Q* + *R* + *M*, all of which were observed.

### 3.2. Model parameters and forecasting

We estimated parameters by fitting the model to actual recorded data from March 1st to April 20th, 2020, and test the model on recorded data thereafter. There are five parameters to estimate. The first is the infectious period (*T* ^*i*^), the period over which an individual can spread the virus (which can occur before developing symptoms) to ceasing to spread the virus because of quarantine, recovery, or death. The second is the basic or initial reproduction number (*R*^0^). The other three parameters include the reduction in community transmission after Australian governments introduced each of the three principal suppression measures, i.e., March 19-21st, March 22-28th, and after March 28, all lagged before coming into effect by the appropriate number of days (see Figure 2).

We fitted the number of recorded cases projected by the model to the number of observed cases using a non-linear least squares technique (Green, 2012) that estimated the parameters by minimizing the sum of the squared distance between the projected and actual values in equation (6):

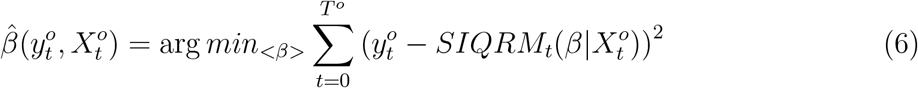

where 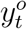 is the observed total number of cases (i.e., recorded cases) at time *t*, which differs from the actual number of infected people; 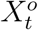 is other observable information at time *t* (e.g., what suppression measures have been introduced up to time *t*); 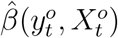 is the (asymptotic mean of) the estimated parameters, conditioning on the observable data and information; *T* ^*°*^ = 51 is the number of days between March 1st and April 20, 2020; 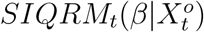 is the total number of recorded cases at time *t* that is projected by the SIQRM model given observable information 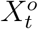 and a set of parameters *β*.

Following Green (2012), the estimated asymptotic variance-covariance (ESV) matrix of the non-linear least square estimators was calculated using equation 7

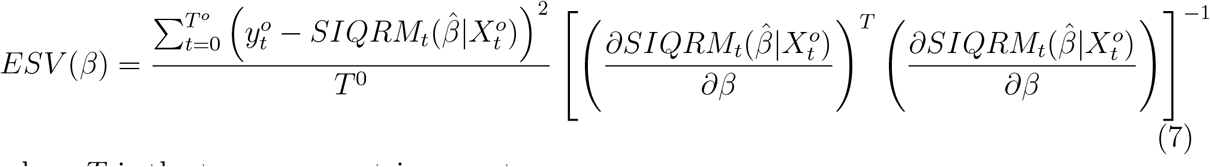

where *T* is the transpose matrix operator.

A computational issue arising from the non-linear least squares technique in that it depends on the starting point of each parameter value, and the optimization process may end up with a local rather than the global minimum. Our response was to use a multi-start optimization algorithm. Thus, we repeatedly solved the least-squares optimization process with 1,000 random starting ‘guess points’ and used the best point to simulate the projection outcomes. These starting ‘guess points’ were randomized within certain ranges, as suggested by parameter values in the literature. For example, the range for randomizing the initial guess for the basic reproduction rate was 1.4-3.25 (Australian Department of Health, 2020c), and the range for the initial guess of the infectious period was between 1-14 days (Lauer et al., 2020). The range for the effectiveness of suppression measures was set between 0 and 1, by the definition of that parameter.

The estimated parameters to forecast the dynamics of the spread of reported COVID-19 was undertaken by randomizing the parameters using their estimated distributions to generate 20,000 simulations with a parallel-processing routine. To test the predictive capacity of the model, we followed a standard procedure that used some part of the available data to fit the model, then projected forward to determine how the model matched the remaining available data. In particular, March 1st (the first recorded death) was considered as time zero, and we used the next 50-day interval (from March 2nd to April 20th, inclusive) to estimate model parameters, or to ‘fit the model’. Data from April 22nd were used to determine model ‘accuracy’. Suppression measures were introduced to account for the period of time (on average) before symptoms appear and, thus, when new cases were recorded in the data.

### 3.3. Model fit

Figure 3 compares the mean-value projections generated by our SIQRM model and the actual data (relevant confidence intervals are provided in the Figure 4). The three panels compare three observable indicators, namely; the total number of recorded cases, the number of active cases recorded, and the number of fatalities. The red line is model output, the blue line is the actual data, which we used to fit the model to April 21st, and the dotted line is actual post-fitted data. The model outcome for peak timing of recorded active cases (roughly April 5th) is in the middle panel and closely matches actual data.

**Figure 3:**
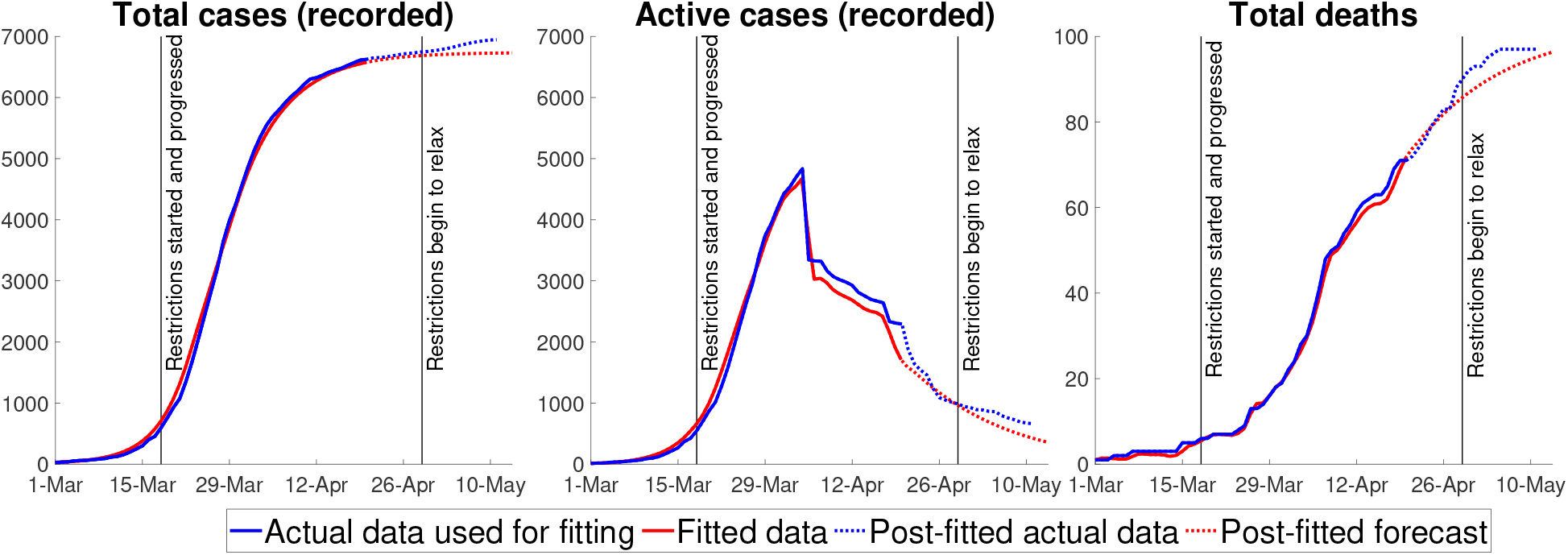
Projected versus Actual Data.

**Figure 4:**
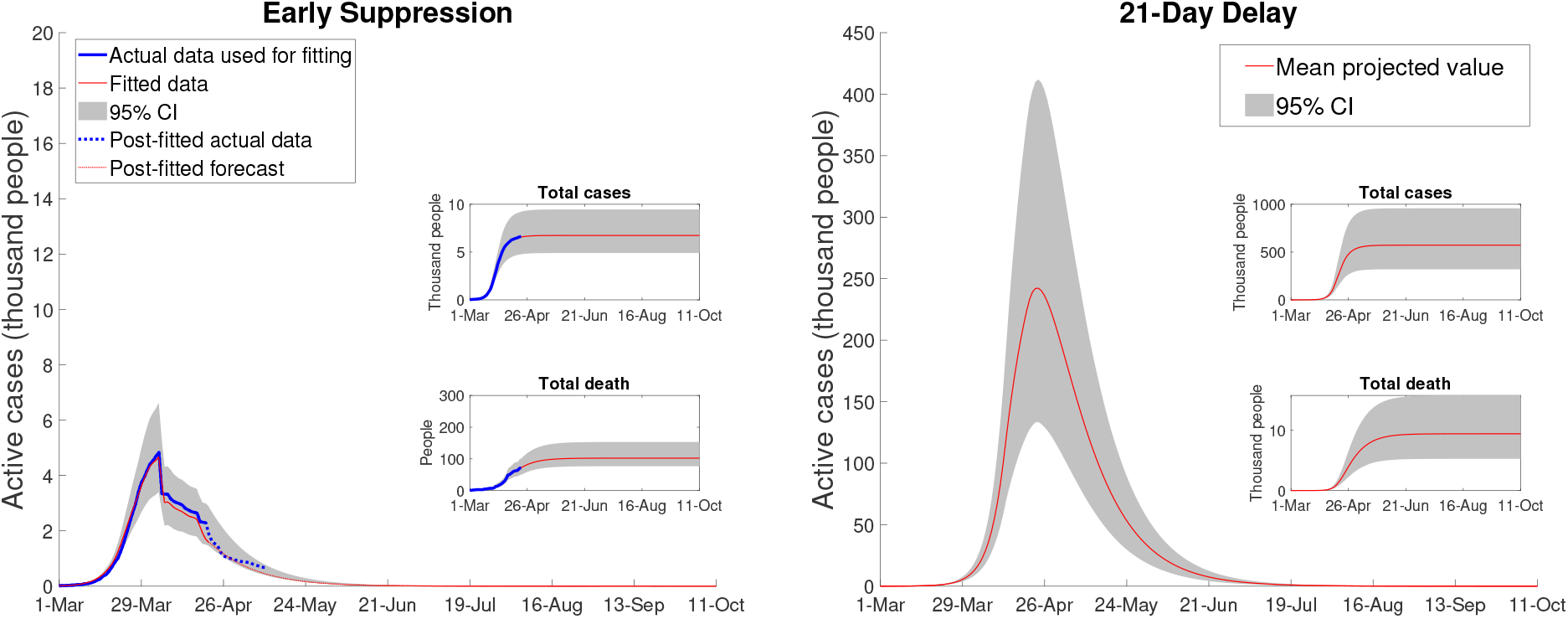
Australian COVID-19 Dynamics: Early and Delayed (21 days) Suppression.

The two key estimated epidemiological parameters used to generate Figure 3, along with their 95% confidence intervals, are (1) average infection period *T* ^*i*^ in days, 7.00 [6.85-7.15] and the (2) basic reproduction rate *R*_0_, 2.48 [2.45 – 2.51]. The effectiveness of the three policy measures, estimated as percentage reductions in transmissions compared to the no suppression scenario or, in effect, (1*−∈*) in the SIQRM model, are: (3) bans on non-essential gatherings 92.19 [83.71-100], (4) non-essential business shutdown 38.14 [35.50-40.77] and (5) maximum two-person gatherings 3.5 [2.97-4.04]. A higher value of the policy parameter ∈, or a lower (1*−* ∈), means a greater effect on reducing the rate of infection. Bans on non-essential gathering, for example, reduced transmission by roughly 8% with the most important suspension measure coming from the limits on the number of people in a gathering.

Our results represent the impacts of a combination of suppression measures, especially non-essential business shut down and limits on gathering, noting that these measures likely reinforce each other. Combined with voluntary social distancing and home isolation, the three suppression measures decreased the extent of the transmission by an estimated (roughly) 96.5%. Our estimates are consistent with surveys by the Australian Bureau of Statistics (ABS), ABS (2020d), in the first week of April showing that 98 per cent of the population was practising social distancing and 88 per cent avoided public spaces and events.

### 3.4. Health care facilities

The requirements for health care facilities (e.g., hospitalization, mechanical, and non-invasive ventilators) are approximated via the following equations:

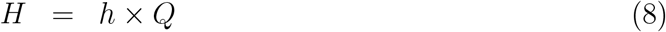

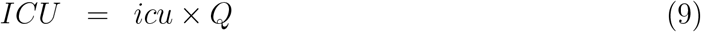

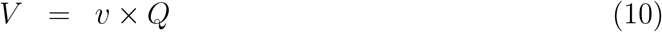

where *H* is the number of hospitalized patients and *h* is the hospitalization ratio (to the number of active cases). We used an estimate from the Australian Department of Health (2020*b*) and Garg, et al. (2020) to specify *h* = 4.6%; *ICU* is the number of patients who require an ICU bed and *icu* is the ratio of those patients to the number of active cases (Science Media Centre, 2020). We specify *icu* = 1.5% using the estimate of Garg, et al. (2020) and Fox et al. (2020); *V* is the number of patients who require a ventilator, and *v* is the ratio of those patients to the number of active cases. The value of *v* is estimated to be 1.43% (Grasselli, et al., 2020).

To determine impacts on patient welfare, we used both a Value of a Statistical Life Year (VSLY) measure and an age-adjusted Value of Statistical Life (a-VSL). We avoided a quality-adjusted life year (QALY) measure because we question its usefulness for cost-effectiveness comparisons among alternative public health responses in a pandemic when the health care budgets vary so greatly among the alternatives. We also note there are identified challenges, including fairness, in the application of QALY measures (Nord et al., 2009).

In the first instance, we estimated welfare losses as the difference between normal life expectancy in Australia and average age at the time of death from COVID-19:

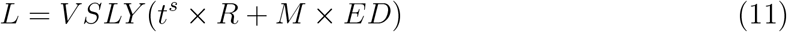

where VSLY is an estimate of the value society places on a year of life, in principle measured by estimating the marginal value or willingness to pay to reduce the risk of death. We use an updated VSLY (Abelson, P., 2007), as applied by the Australian Government for public decision-making, of $213,000, independent of age (Prime Minister and Cabinet, 2019). *ED* in equation (11) is the difference between normal life expectancy and the average age at death of patients who can not recover. We estimated *ED* = 6.9 years, using the average life expectancy in Australia of 82.5 years (ABS, 2020a), (ABS, 2019), and estimate the average age at death from COVID-19 at 75.6 years (Covid19-Data, 2020). The value *t*^*s*^ is the average number of sick days of patients who recover, specified at 18.5 days on average (Covid19-Data, 2020).

Rather than simply averaging, we use a measure of VSL adjusted by age to account for the fact that most of the deaths in Australia from COVID-19 are among the elderly. Of the 103 fatalities as of 31st May, only 5 were less than 60 years of age, 10 were in their 60s, 34 in their 70s, 34 in their 80s and 20 in their 90s. The VSL was obtained from government estimates and given by $4.9 million (Prime Minister and Cabinet, 2019). We scaled this Australian VSL by .70 for those over 70 years of age, following Alberini et al. (2004), who obtained age-adjusted ‘willingness to pay’ measures for reductions in mortality risk using contingent valuation surveys. We assumed that the age distribution is unchanged as the number of deaths varies.

## 4. Results

Figure 4, shows both the effect of the 8-weeks lock-down from early suppression and the effect of an assumed 21-day delay in introducing the full range of suppression measures. Early suppression in Australia has been effective at both ‘flattening and shortening’ the curve. Our model closeley approximates the actual data and estimates the total number of active cases peaks at (roughly) 4,850 cases, with 100 deaths and 6,650 total cases (for a 95% confidence interval of [4,032–12,082] in case load).

With a 21-day delay suppression strategy, the number of active cases peaks at 241,000 [132,000–405,000] and the number of deaths increases to 9,074 [5300–15,360]. The period over which severe suppression measures is extended runs until the third week of July, where there are roughly only 500 active cases remaining. With a 14-day delay the number of fatalities are 2,144 [1,312–3,334] and with a 28-day delay the number of fatalities are 35,374 [19,850–59,070].

Figure 5 shows the case of no suppression, the extreme counterfactual. The total number of cases is nearly 16 million, with a peak of 5.7 million active cases. Fatalities without control, are roughly 260,000 (assuming Australia’s current reported mortality rate, or reported deaths as a fraction of reported cases). Noting the difference in scale in the two panels in Figure 5, with no controls, the projected number of active cases still exceeds 2,100 in August 2020 and COVID-19 is not effectively suppressed until late December 2020. Using the lower fatality ratio of 0.7% as found in Verity et al. (2020), results in roughly 112,000 deaths with the no suppression scenario. We include this case, along with the Australian reported fatality rate, in the economic estimates to follow since although the Australian fatality rate on recorded cases predicts well for the lock-down and delayed suppression cases (with their limited horizons), a different fatality rate may apply for the (out of sample) no suppression case.

**Figure 5:**
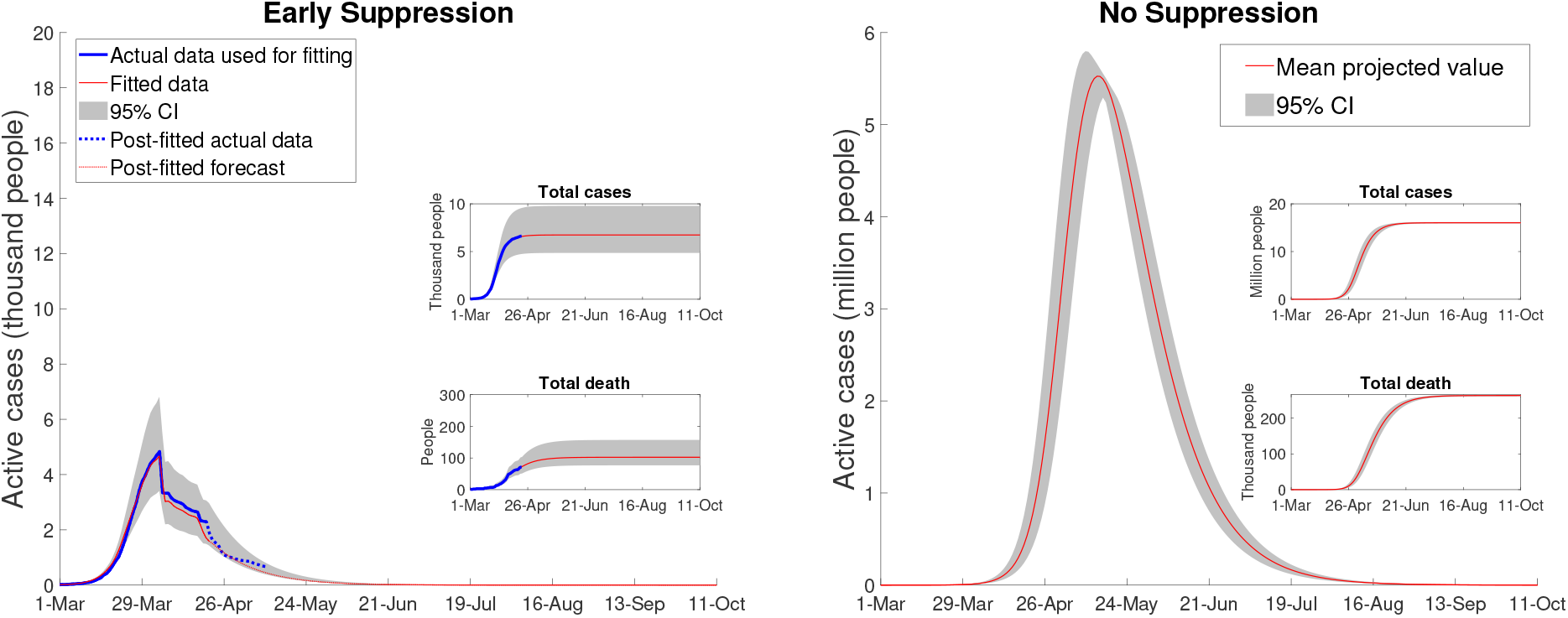
Australian COVID-19 Dynamics: Early and No Suppression.

### 4.1. Public health outcomes

Using the results represented in Figures 4 and 5, we projected the demand for health care facilities for early, delayed and no suppression measures. With early suppression, the number of hospital patients is 220 at its peak, with a 95% confidence interval of [130–380]. The peak number of ICU beds and ventilators are, respectively, about 80 and 70 with early suppression. With no suppression, assuming no capacity limits, the number of hospitalized patients peaks at more than 260,000, of which more than 80,000 would need to be placed in an ICU. This ICU use estimate is nearly 40 times higher than the capacity of Australia’s health care system, with only 2,229-2,378 ICU beds (Australian and New Zealand Intensive Care Society, 2019), (Litton, et al., 2020).

Without an extended lock-down, roughly 70,000 patients would require ventilators at the peak, while there are only around 4,820 machines capable of delivering invasive mechanical ventilation (Litton, et al., 2020). For 21-day delayed suppression, hospital beds peak at 11,100 [6,000–18,640], and ICUs at 3,585 [1,980–6,075], both also exceeding the capacity of the Australian medical care system.

### 4.2. Welfare losses from Covid19 patients

Tables 1 and 2 provide welfare loss estimates of COVID-19 patients using an Australian VSL of $4.9 million, scaled down or adjusted by .70 for those over 70 years of age, and a VSLY of $231,000 AUD. The VSL estimates in Table 1 vary substantially depending on the scenario. Welfare damages are minimized with early suppression. For the case of a fatality rate of 0.7%, is $401.6 billion.

**Table 1:**
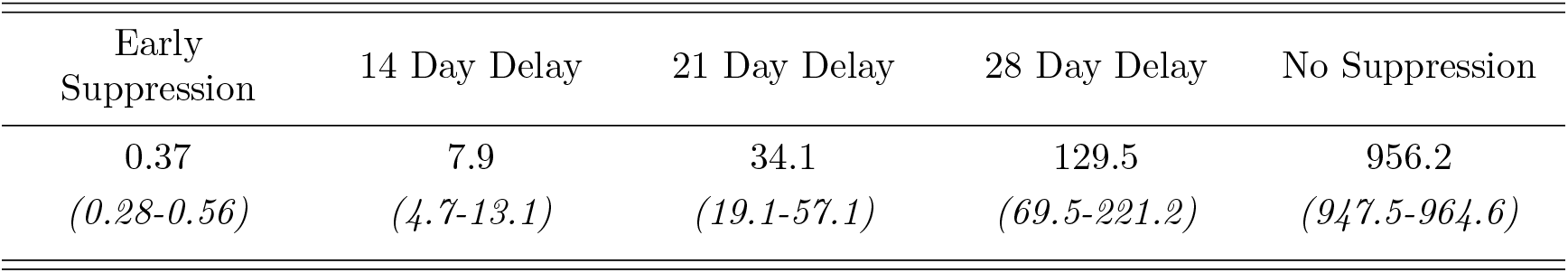
A-VSL Mortality Estimates by Scenario (*$ billion AUD*)

| Early Suppression | 14 Day Delay | 21 Day Delay | 28 Day Delay | No Suppression |
| --- | --- | --- | --- | --- |
| 0.37 | 7.9 | 34.1 | 129.5 | 956.2 |
| (0.28-0.56) | (4.7-13.1) | (19.1-57.1) | (69.5-221.2) | (947.5-964.6) |

**Table 2:** Welfare Losses of Covid-19 Patients: Early versus No Suppression

| | Early Suppression (\$<br>*million*) | No Suppression (\$<br>*billion*) |
| --- | --- | --- |
| Recovered | 80.8<br>[58.5–113.7] | 192.1<br>190.3–193.7 |
| Fatalities | 148.2<br>[111.9–221.4] | 380.8<br>[337.3–383.8] |
| Total | 229<br>[170.4–335] | 572.8<br>[567.6–577.5] |

Table 2, with the VSLY measure, shows that, with early suppression, the total welfare loss is approximately $229 million AUD with a 95% confidence interval of [$170.4–$335] million AUD. Around 30% of this total cost is attributed to those who recover and around 70% of the costs are from fatalities. For the no suppression scenario, the losses are very large, amounting to $572.8 billion [567.7–577.5] AUD, or roughly 2,500 times the loss of early suppression. Losses with the 0.7% fatality rate are $240.6 billion, or 1,048 times larger than early suppression.

### 4.3. Health care costs

Table 3 reports the estimate of hospitalization costs, using average measures of bed costs (IPHA (2020), IPHA (2016)), for early suppression and no suppression, using VSLY. For early suppression, the mean value of the total hospitalization costs is $9.8 million AUD of which $3.9 million is the cost of ICU services and $5.9 million is for occupied hospital beds. For the no suppression scenario, the number of hospitalizations is more than 2,300 times higher and the projected hospitalization costs are at least $23.3 billion AUD, with a 95% confidence interval of $23-$27.3 billion AUD. For the 0.7% fatality rate, total costs are $5.88 billion for the no suppression case, or nearly 1000 times larger. Hospitalization costs do not include pre-hospital costs (e.g., such as visits to the GP or ambulance services) or any costs occurring post-hospitalization (e.g., follow-up expenses, and related health risks from having contracted the virus, which are known to be significant).

**Table 3:** Estimated Hospitalization Costs: Early versus No Suppression

| | Early Suppression (\$<br>*million*) | No Suppression (\$<br>*billion*) |
| --- | --- | --- |
| Total Costs | 9.8<br>(6–18) | 23.3<br>(23–23.7) |
| ICU beds | 3.9<br>(2.4–7.2) | 9.3<br>(9.2–9.5) |
| Hospital beds | 5.9<br>(3.6–10.8) | 14<br>(13.8–14.2) |

### 4.4. Direct economy-wide costs

To measure the direct economy-wide costs from early suppression, we used a survey conducted by the ABS of business activity during the period from March 30th to April 3rd, after the March 21st shut-down of all non-essential businesses, and again in May (ABS, 2020*b*);(ABS, 2020*c*). Some 84 percent of businesses reported that suppression measures affected their activity, with many operating at reduced levels, or not at all. Using the reduction of business activity provided by the ABS across all categories, we apportioned the economic cost per day of suppression measures as *C* by:

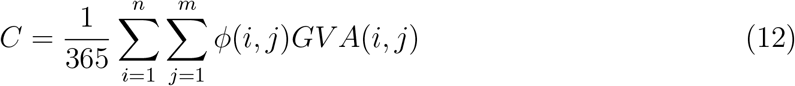

where *ϕ*(*i, j*) is the loss of gross value added in region *i*, sector *j* given the suppression measures and *GV A*(*i, j*) is the gross value of production in 2019 (ABS, 2020e). The value of *ϕ*(*i, j*) was estimated as:

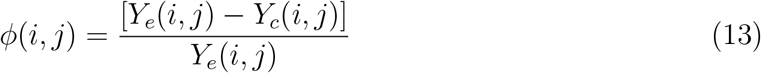

where *Y*_*e*_(*i, j*) and *Y*_*c*_(*i, j*) is the expected and actual level of business before and after the full set of suppression measures were in place for COVID-19, or ∈= 0.965 in our SIQRM model, drawn from the ABS surveys (ABS, 2020b);(ABS, 2020c). It is important to note in these measures that we adjusted the ABS estimates on mining activity following Claughton et al. (2020), and that we account for IMF (2020) forecasts for the 6% fall in global economic activity, projected to occur (with or without the Australian lock-down).

The value of *C* is captured in an aggregated form (across the various ABS tables and adjustments) in Table 4. Total losses are $982.2 million per day, or roughly $6.5 billion per week. In terms of order of magnitude, $982.2 million per day is about a 17% fall in daily GDP (using an annual GDP in 2019 of $1,995 billion AUD). Figure 6 shows the impacts by state and territory in Australia, with NSW the most adversely affected.

**Table 4:** Cost of Control Measures *($ million per day AUD)*.

| Sector/Region | NSW | VIC | QLD | SA | WA | TAS | NT | ACT | Sum |
| --- | --- | --- | --- | --- | --- | --- | --- | --- | --- |
| Mining | 11.9 | 3.2 | 30.5 | 2.3 | 67.9 | 0.7 | 4.3 | 0.0 | 120.9 |
| Manufacturing | 29.2 | 23.4 | 19.0 | 4.9 | 10.3 | 1.8 | 1.1 | 0.4 | 89.9 |
| Electricity, gas, water, waste | 6.9 | 4.5 | 5.3 | 1.8 | 2.6 | 0.9 | 0.3 | 0.3 | 22.5 |
| Construction | 31.7 | 26.2 | 19.3 | 5.7 | 14.4 | 1.5 | 3.3 | 1.8 | 103.9 |
| Wholesale trade | 12.0 | 9.6 | 7.6 | 2.3 | 3.5 | 0.5 | 0.2 | 0.3 | 36.0 |
| Retail trade | 8.4 | 7.0 | 5.2 | 1.8 | 2.7 | 0.6 | 0.3 | 0.5 | 26.5 |
| Accom & food services | 6.5 | 3.9 | 3.0 | 0.9 | 1.6 | 0.3 | 0.1 | 0.7 | 16.9 |
| Transport, postal, warehouse | 10.5 | 9.8 | 8.4 | 2.5 | 6.8 | 0.9 | 0.7 | 0.5 | 40.2 |
| Infor media & telecom | 10.3 | 6.6 | 3.4 | 1.5 | 1.9 | 0.4 | 0.2 | 0.4 | 24.7 |
| Financial & insurance | 45.0 | 28.0 | 13.3 | 4.9 | 6.7 | 1.1 | 0.4 | 0.9 | 100.4 |
| Rental, hiring, real estate | 13.1 | 7.1 | 7.1 | 1.6 | 2.7 | 0.6 | 1.0 | 0.8 | 34.0 |
| Professional, scientific & tech | 34.4 | 24.8 | 13.2 | 4.0 | 8.9 | 0.7 | 0.8 | 2.4 | 89.2 |
| Administrative services | 17.6 | 11.4 | 9.6 | 2.8 | 5.4 | 0.4 | 0.5 | 0.6 | 48.3 |
| Education & training | 26.4 | 21.1 | 14.6 | 5.0 | 7.0 | 1.6 | 0.8 | 4.4 | 81.0 |
| Health care, social assistance | 22.5 | 17.6 | 14.6 | 5.7 | 8.5 | 1.4 | 0.9 | 1.6 | 72.7 |
| Arts & recreation services | 2.7 | 1.8 | 1.1 | 0.3 | 0.5 | 0.1 | 0.1 | 0.4 | 7.0 |
| Other services | 4.5 | 3.7 | 2.8 | 0.8 | 1.6 | 0.2 | 0.1 | 0.2 | 14.0 |
| Sum | 293.6 | 209.6 | 177.8 | 48.8 | 153.2 | 13.8 | 15.1 | 16.3 | **928.2** |

Note: Calculations based on source survey material on business activity from (ABS, 2020*b*);(ABS, 2020*c*); IMF (2020); and Claughton et al. (2020).

**Figure 6:**
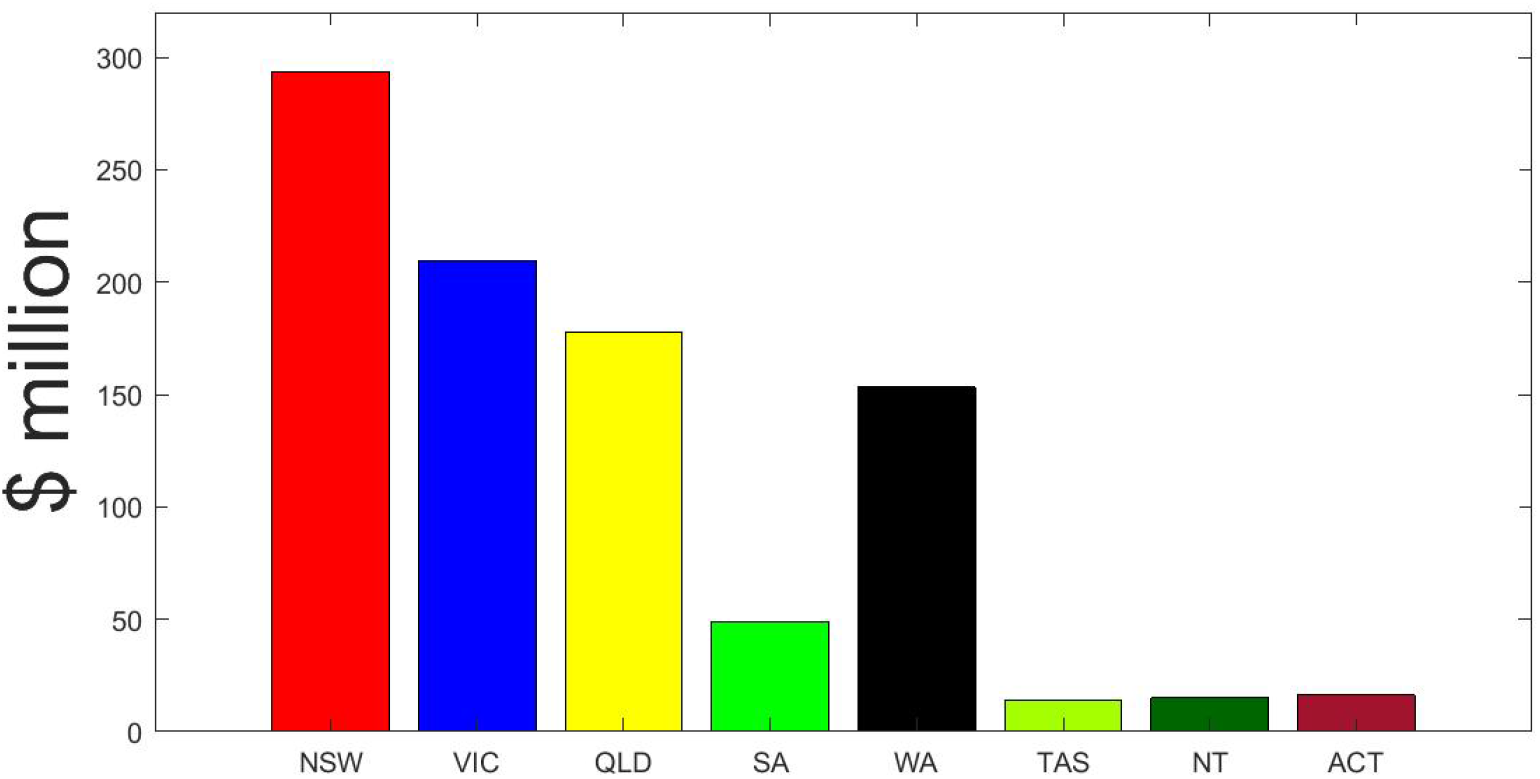
Cost of COVID Control Measures by Region *($ million/day AUD)* ***Note**: NSW: New South Wales; VIC: Victoria; SA: South Australia; WA: Western Australia; TAS: Tasmania; NT: Northern Territory; ACT: Australian Capital Territory.*

### 4.5. Transition costs following a lock-down

Our estimated economic losses from 8 weeks of early suppression measures start from March 30th. These economic losses are the economic cost per day times the number of days under the series of the introduced control measures, or roughly $51.98 billion AUD over the eight-week period, including all international tourism (see Table 4). As suppression measures are relaxed, in the week of May 25th by our simulation, the economy does not ‘snap back’ but transitions to full economic activity over time, taking into account that international travel restrictions will still be in place for some time, or at least until 2021 by our calibration. The speed of transition also depends on how quickly government relaxes controls.

In formal terms, the cost of transition is a generalised function of the extent of control measures:

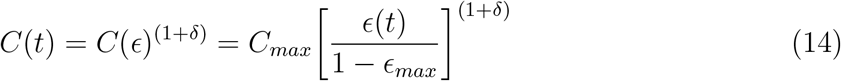

where *C*(*t*) is the cost of control measure at time *t*; (*t*) is the policy measure at time *t*,∈_*max*_ = .965 is the most restrictive policy measure, drawn from the SIQRM model, and *δ* is a cost-policy parameter. We calibrated equation (14) with a knowledge of the cost of the lock-down above at $982.2 million per day and the economic costs of controls conditioned on a estimated value of (*t*) = 0.01, drawn from our SIQRM model, for the international travel ban, or the ‘arrival block’ on China, Iran, South Korea and Italy, prior to more severe domestic control measures being put in place. The losses from the arrival block, *C*_*ab*_, are given by:

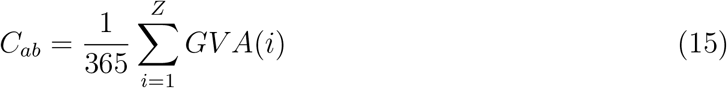

where *GV A*(*i*) is tourism revenue from region *i* and *Z* is number of blocked regions, initially China, Iran, South Korea, and Italy (Tourism of Australia, 2020). The total number of passengers from blocked regions is based on the Bureau of Infrastructure, Transport and Regional Economics (BITRE) (the Department of Infrastructure, Regional Development and Cities of Australia)(BITRE, 2020). Based on BITRE (2020), the ban on travel from the arrival block reduced total inbound passengers, starting in January, by 17.44%. In 2019, 9.3 million international visitors arrived in Australia with a gross value of tourism of $45.4 billion (Tourism of Australia, 2020).Results are summarized in Table 5 and using equation (14) implies:

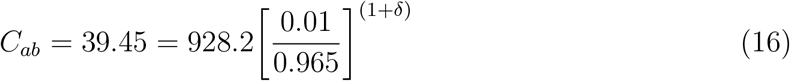

such that

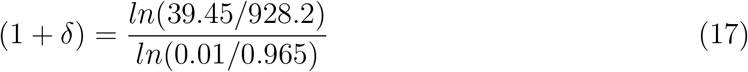

or a value of *δ* = *−*0.309.

**Table 5:**
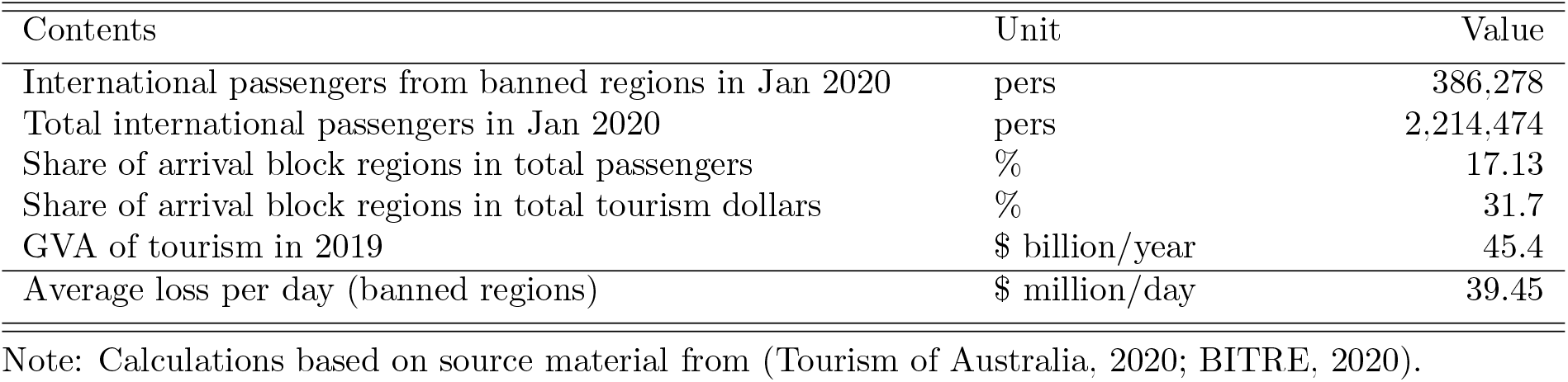
Cost of International Arrival Block

| Contents | Unit | Value |
| --- | --- | --- |
| International passengers from banned regions in Jan 2020 | pers | 386,278 |
| Total international passengers in Jan 2020 | pers | 2,214,474 |
| Share of arrival block regions in total passengers | % | 17.13 |
| Share of arrival block regions in total tourism dollars | % | 31.7 |
| GVA of tourism in 2019 | \$ billion/year | 45.4 |
| Average loss per day (banned regions) | \$ million/day | 39.45 |

Note: Calculations based on source material from (Tourism of Australia, 2020; BITRE, 2020).

Using equation (14), we ‘unwind’ the government control measures and allow for a return of economic activity until ‘full recovery’, so that, after transition, the remaining costs are those largely from losses in international tourism from all countries. If the transition time is as short as 1 month, the domestic economy recovers quickly and the losses in GDP are the smallest. If it takes longer for the economy to recover, modeled as a slower fall in in our formulation, then depending on the period of time over which the transition occurs, economy losses are larger.

### 4.6. Public health and economy costs combined

Table 6 provides a summary of the total economic losses for four different scenarios, by months of transmission for 1, 2, 3 and 4 months duration, drawing on Australian Bureau of Statistics survey data (ABS, 2020*b*);(ABS, 2020*c*) on the cost of the 8-weeks lock-down itself, which we estimate at $51.98 billion. There are two issues to consider; how quickly suppression measures are relaxed and the time it takes for the economy to recover. With early suppression, lock-down measures can be removed more quickly.

**Table 6:** Direct Economic Costs with Early Suppression in $ billions and GDP loss, and Health and Welfare Losses

| | Recovery<br>(months) | Costs (\$ *billion AUD*) | | | Economy<br>Costs (%<br>annual GDP) |
| --- | --- | --- | --- | --- | --- |
|  |  | Lock-down | Recovery | Total | Annual Loss<br>GDP (%) |
| Early Suppression Measures for 8 Weeks from March 30th |  |  |  |  |  |
| Transition 1 | 1 | 51.98 | 14.39 | 66.37 | 3.33 |
| Transition 2 | 2 | 51.98 | 30.39 | 82.37 | 4.13 |
| Transition 3 | 3 | 51.98 | 48.27 | 100.26 | 5.03 |
| Transition 4 | 4 | 51.98 | 68.59 | 120.57 | 6.04 |
| Welfare Losses, Hospitalization Costs and Fatality Equivalents of No Suppression |  |  |  |  |  |
|  |  | Welfare | Hospital | Total | Annual Loss<br>GDP (%) |
| VSLY |  | 572.8 | 23.3 | 596.1 | 29.8 |
| VSLY* |  | 240.0 | 23.3 | 263.3 | 13.1 |
| A-VSL |  | 956.2 |  | 956.2 | 47.9 |
| A-VSL* |  | 401.6 |  | 401.6 | 20.1 |
| Fatality<br>equivalent<br>at %<br>GDP** |  | 30,491 (3.3%) | 37,816 (4.13%) | 46,057 (5.03%) | 55,305 (6.04%) |
| Fatality<br>equivalent<br>at %GDP* |  | 12,808 (3.3%) | 15,882 (4.13%) | 19,343 (5.03%) | 23,228 (6.04%) |

\*VSLY, A-VSL and fatality equivalent measures using the fatality ratio in Verity et al. (2020). \*\*Fatality equivalent is the VSLY-measured number of fatalities under the no suppression scenario that equals the direct economy cost associated with an early 8-weeks lock-down (early suppression) for each % GDP loss (3.33, 4.13, 5.03, and 6.04). N.B. The estimated early suppression model fatalities are 100.

We assume that the cost of transition is roughly half the monthly losses in lock-down in the first month, with costs of transition convex in recovery time in the remaining months. We also note that the cost of the lock-down itself already includes losses in international tourism for all countries, which continue through transition. Given a convex cost of recovery, our simulation gives a range of annualized losses in GDP of early suppression from 3.33% to 6.04% (see Table 6). With no suppression measures, the welfare and hospitalization costs as a percentage of annual GDP range from 13.1% to 47.9%, depending on the welfare measure.

## 5. Discussion

Whether early suppression results in a higher overall economic cost than delayed or no suppression measures requires specification of the severity and duration of the lock-down and reliable estimates in relation to key public health parameters. Using a fit-for-purpose SIQRM model for Australia, we estimated the number of cases (cumulative and active), fatalities, hospitalization costs, welfare losses for COVID-19 patients and the loss in economic activity under early suppression and various delays (14, 21 and 28 days), including the case of no suppression. Total estimated hospitalization costs, welfare losses and number of deaths range from more than 1,000 to 4,000 times larger with no suppression compared to early suppression. Delays in suppression (14 to 28 days, or more) provide no economic gain, but increase fatalities and also lengthen the period over which suppression measures are required before active cases fall below 500.

Direct economy costs of early suppression depend on how quickly the economy recovers following a lock-down. In the fastest (1 month) assumed recovery after lock-down, the total economy-wide costs are about 3% of GDP. With the slowest assumed recovery (4 months), the total direct economy costs are some 6% of GDP. By contrast, the total welfare losses and hospitalization costs range from 12.5% to nearly 48% of GDP with no suppression measures in place. Our results indicate that the total economic costs of no suppression are (roughly) between 4 and over 8 times larger than early suppression (see Table 6).

We provide a number of caveats to our work. First, our VSLY measure underestimate the welfare losses associated with no suppression measures. This is because we only have limited information to benchmark on average life expectancy, noting that many of those who die of COVID-19 are older than 82.5 years. In our view, people in this older age bracket would have a both a willingness and ability to pay for additional life. Second, in the case of no suppression measures, we assume no constraints on hospital and ICU capacity noting that a capacity constraint would likely increase fatalities, and thus welfare losses, should such a constraint be exceeded. Third, for the no suppression scenario, we do not explicitly model the effects of voluntary physical distancing or a delayed lock-down that could occur with substantial community infections and deaths. Instead, we estimate the welfare losses (VSLY of fatalities) equivalent to the GDP costs of an 8-weeks lock-down (see Table 6). We find that VSLY welfare losses of fatalities, equivalent to GDP losses from an early 8-weeks lock-down, would have resulted in more than 12,500–30,000 Australian deaths depending on the fatality rate – fatalities comparable to countries (e.g., United Kingdom) that implemented a delayed lock-down. In other words, for early suppression *not* to be the preferred response to COVID-19, requires that Australians prefer more than 12,500-30,000 deaths from no suppression measures (actual Australian deaths are 102 as of 31 May 2020) than the direct economic costs associated with an early 8-weeks lock-down. Fourth, data limitations preclude us from estimating indirect social costs of unemployment with early or delayed suppression (e.g., additional suicides, domestic violence and alcoholism), but note that many Australian workers have retained employment through a government subsidised and temporary ‘JobKeeper’ scheme that began on March 30th, 2020.

Notwithstanding our caveats, from both a public health (mortality and morbidity) and direct economic costs (including public health costs) perspective, Australia’s early suppression measures that began in March 2020, and which we assume would have continued until the end of May 2020 (suppression measures began to be progressively relaxed from early May), generate a very large economic payoff relative to an alternatives of delayed or no suppression measures. At least for Australia, the disease (COVID-19) is, indeed, much more costly than an early ‘cure’ (lock-down).

Our findings provide robust evidence that ‘go hard, go early’ suppression, at least in a high-income country like Australia, is the preferred approach from both a public health and an economy perspective. By comparison, some high-income countries, such as the United States of America (USA), adopted a delayed lock-down when infection rates were much higher than when Australia began its lock-down. Our model results suggest that if the USA, and possibly other high-income countries, had imposed effective suppression measures earlier they may have reduced both their public health costs (including lower fatalities per 1,000 people) and economy costs.

## Data Availability

Data and MatLab source code is available on request.

## Author Contributions

TK and RQG conceived the approach. TK, TNC and LC were responsible for data analysis, initial model construction, and statistical estimations. TK, RQG, TNC, and LC revised the model. TK, RQG, TNC, LC and JC interpreted the results. RQG, TK, TNC, LC and JC wrote the manuscript.

## Acknowledgments

The authors are grateful for helpful comments and suggestions provided by Emily Banks, John Baumgartner, Nathaniel Bloomfield, John Parslow and Andrew Robinson on earlier versions of this manuscript.

## References

Abelson, P. (2007), ‘Establishing a Monetary Value for Lives Saved: Issues and Controversies’, Working Papers in Cost benefit Analysis WP 2008-2, Department of Finance and Deregulation. Accessed 26 April 2020. Available at www.pmc.gov.au.

ABS (2017), ‘Australian Demographic Statistics’, ABS-3101.0-June 2017. Available at www.abs.gov.au.

ABS (2019), ‘Life Tables, States, Territories and Australia, 2016-2018’, ABS 3302. Available at www.abs.gov.au.

ABS (2020a), ‘Births, Australia in 2018’, ABS Stat. Available at www.stat.data.abs.gov.au.

ABS (2020b), ‘Business Indicators, Business Impacts of COVID-19’, ABS-5676.0.55.003. Reported on 30 March. Accessed on 7 April 2020. Available at www.abs.gov.au.

ABS (2020c), ‘Business Indicators, Business Impacts of COVID-19’, ABS-5676.0.55.003. Reported on 4 May. Access on 6 May 2020. Available at www.abs.gov.au.

ABS (2020d), ‘Household Impacts of COVID-18 Survey, 4940.0’, Available at: https://www.abs.gov.au/.

ABS (2020e), ‘Statistics of Australia’, Website. Available at: https://www.abs.gov.au/.

Alberini, A., Cropper, M., Krupnick, A. and Simon, N. B. (2004), ‘Does the value of a statistical life vary with age and health status? evidence from the us and canada’, Journal of Environmental Economics and Management 48 (1), 769–792.

Australian and New Zealand Intensive Care Society (2019), ‘Centre for Outcome and Resource Evaluation - 2018 Report’, ANZICS 2018. Available at www.anzics.com.au.

Australian Department of Health (2020a), ‘Impact of COVID-19 in Australia – ensuring the health system can respond ‘, Australian Department of Health: Impact of COVID-19. Available at www.health.gov.au.

Australian Department of Health (2020b), ‘IMPACT OF COVID-19: Theoretical modelling of how the health system can respond’, Australian Department of Health: Impact of COVID-19. Available at www.health.gov.au.

Australian Department of Health (2020c), ‘Novel coronavirus (COVID-19): Information for Clinicians - Frequently Asked Questions’, Australian Department of Health: Novel coronavirus (COVID-19). Available at www.health.gov.au.

Australian Institute of Health and Welfare (2019), ‘Deaths in Australia’, Australian Institute of Health and Welfare: Report and Data. Available at www.aihw.gov.au.

BITRE (2020), ‘Time Series Statistics of International Airline Activities’, Report. Available at https://www.bitre.gov.au.

Claughton, D., Fowler, C. and Fitzgerald, D. (2020), ‘Mining exploration and service companies hit by coronavirus restrictions’, ABC Rural. Updated 7 April. Access on 6 May 2020. Available at /http://www.abc.net.au.

Covid19-Data (2020), ‘Coronavirus (COVID-19) in Australia’, Website. Available at: https://www.covid19data.com.au/transmission-sources.

Fox, G., Trauer, J. and McBryde, E. (2020), ‘Modelling the impact of COVID-19 upon intensive care services in New South Wales’, The Medical Journal of Australia - Upcoming Publication, Preprint only 30 March 2020.

Frijters, P. (2020), ‘The Corona Dilemma’, Club Troppo. Accessed 6 May 2020. Available at http://clubtroppo.com.au hrefhttp://clubtroppo.com.au/2020/03/21/the-corona-dilemma/.

Garg, et al. (2020), ‘Hospitalization Rates and Characteristics of Patients Hospitalized with Laboratory-Confirmed Coronavirus Disease 2019 — COVID-19’, Morbidity and Mortality Weekly Report. Available at www.cdc.gov.

Grasselli, et al. (2020), ‘Baseline Characteristics and Out-comes of 1591 Patients Infected With SARS-CoV-2 Admitted to ICUs of the Lombardy Region, Italy’, JAMA 323 (16), 1574–1581. URL: https://jamanetwork.com/on05/01/2020

Green, W. (2012), Econometric Analysis, 7th Edition, Pearson Education.

IMF (2020), ‘World Economic Outlook’, WEO Report. Available at http://www.imf.org.

IPHA (2016), ‘National Hospital Cost Data Collection’, Round 18 Report. Available at www.ihpa.gov.au.

IPHA (2020), ‘National Hospital Cost Data Collection’, Round 22 Report. Available at http://www.ihpa.gov.au.

Johns Hopkins University (2020), ‘Coronavirus COVID-19-Global Cases by the Center for Systems Science and Engineering (CSSE)’, Website. Available at www.arcgis.com.

Lauer, S. A., Grantz, K. H., Bi, Q., Jones, F. K., Zheng, Q., Meredith, H. R., Azman, A. S., Reich, N. G. and Lessler, J. (2020), ‘The Incubation Period of Coronavirus Disease 2019 (COVID-19) From Publicly Reported Confirmed Cases: Estimation and Application’, Annals of Internal Medicine. URL: https://doi.org/10.7326/M20-0504

Litton, et al. (2020), ‘Surge capacity of Australian intensive care units associated with COVID-19 admissions’, Medical Journal of Australia. Preprint only-Version 2-April 2020. Available at www.mja.com.au.

Nord, E., Daniels, N. and Kamlet, M. (2009), ‘QALYs: some challenges’, Value in Health 12, S10–S15.

Prime Minister and Cabinet (2019), ‘Best Practice Regulation Guidance Note Value of statistical life’, Primer Minister and Cabinet. Access on 10 May 2020. Available at www.pmc.gov.au.

Science Media Centre (2020), ‘Expert comment on ICU recovery data on COVID-19’, Science Media Centre April 2020. Available at www.sciencemediacentre.org.

Singer, P. and Plant, M. (2020), ‘When Will the Pandemic Cure Be Worse Than the Disease?’, Project Syndicate. The World’s Opinion Page. Accessed 6 May 2020. Available at https://www.project-syndicate.org/.

Tourism of Australia (2020), ‘The Economic Importance of Tourism’, Report. Available at www.tourism.australia.com.

van den Broek-Altenburg, E. and Atherly, A. (2020), ‘Economic Cost of Flattening the Curve’, The Incidental Economist - The Health Services Research Blog. Accessed 6 May 2020. Available at https://theincidentaleconomist.

Verity, R., Okell, L. C., Dorigatti, I., Winskill, P., Whittaker, C., Imai, N., Cuomo-Dannenburg, G., Thompson, H., Walker, P. G., Fu, H. et al.. (2020), ‘Estimates of the severity of coronavirus disease 2019: a model-based analysis’, The Lancet infectious diseases.

Wu, J., Leung, K. and Leung, M. (2020), ‘Nowcasting and forecasting the potential domestic and international spread of the 2019-nCoV outbreak originating in Wuhan, China: a modelling study’, Lancet Public Health 395, 689–697.

